# Effectiveness of Systemic Treatments in Patients with Unresectable, Advanced, or Recurrent Soft Tissue Sarcomas Previously Treated with Anthracycline-Based Therapy: A Systematic Review and Network Meta-Analysis

**DOI:** 10.1101/2025.09.28.25336821

**Authors:** Yosuke Nakano, Satoko Zenitani, Makoto Endo, Takahiro Mihara

**Affiliations:** Department of Health Data Science, Yokohama City University Graduate School of Data Science, Yokohama, Japan; Department of Thoracic Oncology, National Cancer Center Hospital, Tokyo, Japan; Department of Orthopaedic Surgery, Kyushu University Hospital, Fukuoka, Japan

**Author notes:** Corresponding Author: Yosuke Nakano, Contact. Registration: In accordance with the guidelines, our systematic review protocol was registered with the International Prospective Register of Systematic Reviews (PROSPERO) on 28 September 2025 (registration number CRD420251156600). Funding/ Support: This research did not receive any specific grants from funding agencies in the public, commercial, or not-for-profit sectors. Conflict of interest statement: None declared.

**Keywords:** Soft Tissue Sarcoma, Second-line Therapy, Salvage Therapy, Network Meta-Analysis, Randomized Controlled Trials

## Abstract

**Introduction:** Soft tissue sarcomas (STS) are rare malignancies with diverse histological subtypes and an incidence of approximately 40 cases per million annually. Although several agents, including pazopanib, trabectedin, and eribulin, have been approved in Japan since 2012 for patients previously treated with anthracyclines, clinical guidelines provide no consistent recommendations regarding second-line regimens. Given the lack of head-to-head evidence, treatment selection largely depends on clinician discretion. This study aims to evaluate the comparative efficacy of second-line or later pharmacological regimens for unresectable, advanced, or recurrent STS through a systematic review and network meta-analysis (NMA).

**Methods and analysis:** Randomized controlled trials (RCTs), including multi-arm designs, will be identified through comprehensive searches of MEDLINE (via PubMed), Embase (via ProQuest), CENTRAL, WHO ICTRP, and ClinicalTrials.gov from inception to the search date, without language restrictions. Two reviewers will independently screen records, extract data, and assess risk of bias using the Cochrane RoB 2 tool; discrepancies will be resolved by a third reviewer. Pairwise meta-analyses will be performed using random-effects models, followed by a frequentist NMA with between-study variance estimated by REML. Consistency will be assessed using design-by-treatment interaction models (global) and node-splitting methods (local). Treatment ranking will be evaluated using P-scores or SUCRA. Certainty of evidence for each outcome will be assessed using CINeMA in line with the GRADE framework.

**Support/ Funding:** This research did not receive any specific grants from funding agencies in the public, commercial, or not-for-profit sectors.

**Ethics and dissemination:** This systematic review and network meta-analysis will be conducted using data extracted from previously published studies and trial registries. As no individual patient data will be collected or used, ethical approval is not required.

The findings of this review will be disseminated through publication in a peer-reviewed journal and presentation at relevant scientific conferences. The results will also be shared with clinicians, policymakers, and researchers to inform clinical practice and future research.

The final dataset and analytic code may be made available in an open-access repository to ensure transparency and reproducibility.

**PROSPERO Registration number:** CRD420251156600

## 1. Background

Soft tissue sarcomas (STS) are rare malignancies comprising numerous histological subtypes, including liposarcoma and synovial sarcoma, with an incidence of approximately 40 cases per million population annually. In Japan, the Nationwide Bone and Soft Tissue Tumor Registry has been conducted since the 1960s; between 2006 and 2015, 47,852 soft tissue tumors were registered, of which 12,608 were malignant. The five most common subtypes were liposarcoma (4,868 cases), undifferentiated pleomorphic sarcoma (2,584 cases), myxofibrosarcoma (959 cases), leiomyosarcoma (944 cases), and synovial sarcoma (690 cases) (1).

Several new agents have been approved in Japan for previously treated STS since 2012, including pazopanib, trabectedin, and eribulin, all based on trials enrolling patients with prior anthracycline therapy. Although an investigator-initiated comparative study is ongoing domestically, no results have been reported as of August 2025 (2).

The Japanese Clinical Practice Guideline for Soft Tissue Tumors 2020 suggests systemic therapy for unresectable, advanced, or recurrent STS (recommendation grade 2, level C), with doxorubicin monotherapy recommended as first-line (grade 1, level B). Combination therapy with doxorubicin and ifosfamide may be considered when tumor shrinkage is clinically required. For second-line therapy, however, the guideline only notes that the choice depends on histological subtype and treatment goal, without specifying regimens (1). The NCCN Guidelines similarly recommend anthracycline-based regimens first-line, while listing pazopanib, eribulin, trabectedin, and gemcitabine plus docetaxel as category 2A options in subsequent lines (3).

Although certain histologies have drug-specific recommendations, no guideline clearly indicates preferred regimens for second-line STS. In Japan, despite the availability of pazopanib, trabectedin, and eribulin, treatment selection largely depends on the discretion of individual clinicians (1–3).

The rationale for this study is to evaluate the comparative efficacy of pazopanib, trabectedin, eribulin, and other regimens used after anthracycline therapy through a systematic review and network meta-analysis, thereby providing evidence to support clinical decision-making.

## 2. Objectives

To identify effective second-line or later pharmacological treatment regimens from the perspective of disease control in patients with unresectable, advanced, or recurrent soft tissue sarcoma previously treated with anthracycline-based therapy.

### 2.1. Primary Objective

To determine the treatment regimen that contributes most to improving overall survival (OS).

### 2.2. Secondary Objectives

#### 2.2.1. Progression-Free Survival (PFS)

To determine the treatment regimen that contributes most to prolonging PFS.

#### 2.2.2. Objective Response Rate (ORR)

To identify the treatment regimen associated with the highest ORR.

#### 2.2.3. Histology-Specific OS

To determine the treatment regimen that contributes most to improving OS within each histological subtype. Subtypes will be classified into up to five groups, consisting of the most common histologies independently reported across included trials.

#### 2.2.4. Histology-Specific PFS

To determine the treatment regimen that contributes most to prolonging PFS within each histological subtype, following the same classification as in 2.2.3.

#### 2.2.5. Histology-Specific ORR

To identify the treatment regimen associated with the highest ORR within each histological subtype, following the same classification as in 2.2.3.

#### 2.2.6. Treatment Discontinuation Rate

To evaluate the discontinuation rate for each treatment regimen, regardless of the reason for discontinuation. Regimens resumed after a temporary pause will not be considered discontinued.

## 3. PICO

### 3.1. Participants

- Patients with unresectable, advanced, or recurrent soft tissue sarcoma
- History of prior anthracycline-based chemotherapy as first-line treatment
- Aged 15 years or older

### 3.2. Interventions

- Any pharmacological treatment, including chemotherapy, molecular targeted therapy, or immune checkpoint inhibitors

### 3.3. Comparators

- Each intervention compared against other interventions

### 3.4. Outcomes

- Outcomes listed in Section 2 (Objectives): OS, PFS, ORR, histology-specific OS, histology-specific PFS, histology-specific ORR, treatment discontinuation rate, etc.

### 3.5. Exclusion Criteria

- Trials including patients with osteosarcoma*, chondrosarcoma*, plasmacytoma, chordoma, Ewing sarcoma, rhabdomyosarcoma, Kaposi’s sarcoma, gastrointestinal stromal tumor (GIST), or carcinosarcoma (*except for extraskeletal osteosarcoma and extraskeletal myxoid chondrosarcoma, which will be included)
- Trials in which non-pharmacological therapies (e.g., hyperthermia, radiotherapy) are included in the treatment regimen
- Trials in which molecular targeted therapies based on molecular diagnosis (e.g., ALK inhibitors, NTRK inhibitors) are included in the treatment regimen
- Trials in which different routes, schedules, or doses are compared within the same drug regimen

## 4. METHODS

This protocol was developed in accordance with international reporting guidelines for systematic reviews and network meta-analyses, specifically the PRISMA statement (4,5), PRISMA-P (6), and PRISMA-NMA (7).

### 4.1. Types of studies

This review will include randomized controlled trials (RCTs), including both two-arm and multi-arm trials.

Blinding (e.g., open-label, double-blind) will not be considered as an eligibility criterion. Cluster RCTs will be included only if statistical adjustments for clustering are reported or can be derived.

Crossover trials will be included only if data prior to the crossover are available.

Non-randomized trials, observational studies, single-arm trials, case reports, and case series will be excluded.

Conference abstracts will be included only if they provide sufficient methodological information and outcome data; otherwise, they will be excluded but may be used for citation tracking.

No language restrictions will be applied. Where possible, automatic translation will be used for inclusion; if exclusion occurs due to resource limitations, this will be documented.

### 4.2. Search methods for identification of studies

We will search the following databases from inception to the date of the search, without language restrictions:

- MEDLINE (via PubMed)
- Embase (via ProQuest)
- Cochrane Central Register of Controlled Trials (CENTRAL)
- WHO International Clinical Trials Registry Platform (ICTRP)
- ClinicalTrials.gov

Search strategies will be developed using a combination of controlled vocabulary terms (e.g., MeSH, Emtree) and free-text keywords. The PubMed strategy will be adapted for use in each database. Full search strategies for all databases will be provided in the Appendix.

For unpublished or ongoing studies identified through trial registries, we will contact the principal investigators or study sponsors to obtain additional information where possible.

### 4.3. Selection of studies

Two reviewers (YN and SZ) will independently screen titles and abstracts and subsequently assess full texts for eligibility. Any disagreements will be resolved through discussion or, if necessary, adjudication by a third reviewer (ME).

The first stage of screening will assess eligibility based on titles and abstracts, followed by a second stage involving full-text review. Reasons for exclusion at the full-text screening stage will be documented.

Reference management will be conducted using software such as EndNote or Paperpile, and screening will be performed using tools such as Rayyan. The final library will be consolidated and stored in formats such as Research Information Systems (RIS) or National Library of Medicine Bibliographic (NBIB).

Before formal screening, a pilot test will be conducted using a randomly selected subset of studies.

### 4.4. Data extraction and management

Two reviewers (YN and SZ) will independently extract data using a standardized form. The predefined items to be extracted are as follows:

- Trial identifiers (first author, year of publication, country, trial ID, funding source, etc.)
- Study design (parallel-group, multi-arm, blinding, etc.)
- Participant characteristics (median age, sex, ECOG performance status, histological subtype, prior treatments, etc.)
- Intervention details (drug name, regimen, treatment duration, etc.)
- Outcome data (hazard ratios, number of events, follow-up duration, etc.)

In cases where multiple reports originate from the same study, data will be handled as follows:

For studies with multiple publications (e.g., interim and final reports), time-to-event outcomes will be extracted from the report with the longest follow-up, while additional outcomes not included in the final report may be taken from other publications. Outcome-specific source references will be recorded.

Data will be managed using spreadsheet software such as Microsoft Excel. The extraction form will be pilot tested on a small sample of studies prior to full data extraction and refined as necessary.

For missing data, we will attempt to contact the study authors. If the information cannot be obtained, the study will be excluded from the relevant outcome analysis, or alternative approaches such as reconstruction from Kaplan–Meier curves will be considered. Sensitivity analyses will be conducted to assess the impact of missing data.

If additional exploratory items are identified during the review process, they will be recorded in the protocol and handled separately from the predefined items.

### 4.5. Assessment of risk of bias

Risk of bias will be assessed for each outcome using the Cochrane Risk of Bias 2 (RoB 2) tool (8). Two reviewers will perform the assessment independently, and any disagreements will be resolved by discussion or, if necessary, adjudication by a third reviewer (ME).

In addition, bias specific to network meta-analysis due to missing evidence will be assessed using the Risk of Bias due to Missing Evidence in Network Meta-analysis (ROB-MEN) tool (9). The risk of missing evidence will first be examined for each pairwise comparison and then synthesized across the entire network.

All assessments will be conducted independently by two reviewers for each outcome, with discrepancies resolved through consultation with a third reviewer (ME).

### 4.6. Measures of treatment effect

Depending on the type of outcome, the following effect measures will be used. In all analyses, effect estimates will be oriented so that greater efficacy or lower risk is expressed as a positive effect.

#### 4.6.1. Time-to-event outcomes (OS, PFS, etc.)

For time-to-event outcomes, the effect measure will be the hazard ratio (HR) with 95% confidence intervals (95% CI). Reported HRs from each trial will be used when available. If not reported, HRs will be reconstructed from Kaplan–Meier curves, number of events, follow-up periods, or other available data using established methods (e.g., Parmar method, Tierney method).

Analyses will, in principle, be based on the intention-to-treat (ITT) population.

#### 4.6.2. Binary outcomes (ORR, treatment discontinuation, etc.)

For binary outcomes, the effect measure will be the risk ratio (RR) with 95% confidence intervals (95% CI). Sensitivity analyses using odds ratios (ORs) will be conducted as necessary.

The denominator will be based on the intention-to-treat (ITT) population.

### 4.7. Data synthesis

We will first perform pairwise meta-analyses for each intervention comparison to evaluate effect estimates and heterogeneity derived from direct comparisons.

Subsequently, a network meta-analysis (NMA) will be conducted to integrate evidence across the entire network (10).

#### 4.7.1. Pairwise meta-analysis

For each intervention comparison, pooled estimates will be calculated using a random-effects model. Depending on the outcome, effect measures will include hazard ratios (HRs), risk ratios (RRs), odds ratios (ORs), or mean differences (MDs)/standardized mean differences (SMDs), with 95% confidence intervals (95% CIs).

Statistical heterogeneity will be assessed using the I² statistic and τ², and prediction intervals will also be reported. We will interpret I² values of approximately 25%, 50%, and 75% as representing low, moderate, and high heterogeneity, respectively (10). Clinical and methodological heterogeneity will additionally be considered.

#### 4.7.2. Network meta-analysis

A frequentist network meta-analysis (NMA) based on a random-effects model will be conducted using R packages such as netmeta and metafor. Between-study variance (τ²) will be estimated using restricted maximum likelihood (REML). For multi-arm trials, correlations will be accounted for using a variance–covariance matrix derived from Rücker’s method (11).

Consistency across the entire network will be assessed globally using the design-by-treatment interaction test and locally using the node-splitting method. Treatment ranking will be evaluated using P-scores or the surface under the cumulative ranking curve (SUCRA) (12,13).

#### 4.7.3. Publication bias and small-study effects

To assess publication bias and small-study effects, we will construct comparison-adjusted funnel plots by re-expressing the entire network relative to a common comparator, and test for asymmetry using Egger’s regression (10). Begg’s test will also be used as a sensitivity analysis if necessary. In addition, we will cross-check against clinical trial registries (WHO ICTRP, ClinicalTrials.gov) to identify unpublished studies.

### 4.8. Subgroup and sensitivity analysis

The following prespecified subgroup analyses will be conducted:

- By histological subtype (e.g., liposarcoma, leiomyosarcoma, synovial sarcoma)
- By patient characteristics (e.g., age group, prior exposure to ifosfamide)

Subgroup analyses will be performed only when sufficient data are available.

Additionally, if adequate data are reported at the trial level, exploratory meta-regression analyses will be conducted to examine potential effect modifiers such as histological distribution, line of therapy, and performance status.

Sensitivity analyses will be performed under the following conditions:

- Excluding trials judged to be at high risk of bias
- Excluding trials reporting time-to-event outcomes solely from Kaplan–Meier curves
- Restricting treatment discontinuation analysis to discontinuation due to toxicity, where data are available

As a sensitivity analysis, treatment nodes may be redefined as follows: agents specifically approved for STS (e.g., pazopanib, trabectedin, eribulin) will be retained as independent nodes; combination regimens (e.g., gemcitabine plus docetaxel) will also be retained as independent nodes; and remaining monotherapy agents without STS-specific approval (e.g., dacarbazine, ifosfamide) will be consolidated into a single “traditional chemotherapy” node.

### 4.9. Assessment of transitivity and inconsistency

To assess transitivity, we will examine the distribution of potential effect modifiers (e.g., histological distribution, line of therapy, performance status) across comparisons. If substantial differences in these distributions are judged to be clinically inappropriate, the corresponding comparisons will be excluded or combined. Trials enrolling patients with a single histological subtype will be included in histology-specific analyses only and excluded from the primary analyses of the overall population.

Assessment of inconsistency will be conducted as follows:

- Global assessment: using the design-by-treatment interaction model
- Local assessment: using the node-splitting method

If inconsistency is identified, additional sensitivity or subgroup analyses will be performed to explore potential sources.

### 4.10. Assessment of the certainty of evidence

We will assess the certainty of evidence for each outcome using CINeMA (Confidence in Network Meta-Analysis) (14). The following six domains will be evaluated:

1. Within-study bias (bias within individual trials)
2. Reporting bias (including publication and selective reporting bias across studies)
3. Indirectness
4. Imprecision
5. Heterogeneity
6. Incoherence

In addition, following the extension of the GRADE approach to network meta-analysis, results will be presented as Summary of Findings tables for each outcome.

## Supporting information

Version history_20260719

## Data Availability

All data referred to in this protocol are derived from previously published articles and publicly accessible trial registries (e.g., MEDLINE, Embase, CENTRAL, WHO ICTRP, ClinicalTrials.gov).
No individual patient-level data will be collected. All data extracted and analyzed in this study will be fully contained in the published manuscript or its supplementary materials.

## 5. Appendix

### 5.1. Search strategy for MEDLINE & PubMed Central

| Search | Query |
| --- | --- |
| #1 | "Sarcoma" [MeSH] OR "Soft Tissue Neoplasms" [MeSH] OR "Neoplasms, Connective and Soft Tissue" [MeSH] OR "Soft Tissue Sarcoma" [tiab] OR "STS" [tiab] OR "Sarcoma" [tiab] |
| #2 | "Neoplasm Recurrence, Local" [MeSH] OR "Neoplasm Metastasis" [MeSH] OR "Neoplasm, Residual" [MeSH] OR "unresectable" [tiab] OR "advanced" [tiab] OR "recurrent" [tiab] OR "metastatic" [tiab] OR "locally advanced" [tiab] |
| #3 | #1 and #2 |
| #4 | "drug therapy" [Subheading] OR "chemotherapy" [tiab] OR "systemic therapy" [tiab] OR "second-line" [tiab] OR "salvage therapy" [tiab] OR "targeted therapy" [tiab] OR "pharmacotherapy" [tiab] OR "immune checkpoint inhibitor*" [tiab] OR "PD-1 inhibitor*" [tiab] OR "PD-L1 inhibitor*" [tiab] OR "CTLA-4 inhibitor*" [tiab] |
| #5 | "pazopanib" [tiab] OR "trabectedin" [tiab] OR "eribulin" [tiab] OR "gemcitabine" [tiab] OR "docetaxel" [tiab] OR "doxorubicin" [tiab] OR "ifosfamide" [tiab] OR "nivolumab" [tiab] OR "pembrolizumab" [tiab] OR "atezolizumab" [tiab] OR "avelumab" [tiab] OR "durvalumab" [tiab] OR "ipilimumab" [tiab] OR "olaratumab" [tiab] OR "brigimadlin" [tiab] |
| #6 | #4 and #5 |
| #7 | (randomized controlled trial [pt] OR controlled clinical trial [pt] OR randomized [tiab] OR placebo [tiab] OR drug therapy [sh] OR randomly [tiab] OR trial [tiab] OR groups [tiab]) NOT (animals [mh] NOT humans [mh]) |
| #8 | #3 and #6 and #7 |

### 5.2. Search strategy for Cochrane CENTRAL

| Search | Query |
| --- | --- |
| #1 | Sarcoma:ti, ab, kw OR "Soft Tissue Neoplasms":ti, ab, kw OR "Neoplasms, Connective and Soft Tissue":ti, ab, kw OR "Soft Tissue Sarcoma":ti, ab, kw OR STS:ti, ab, kw |
| #2 | "Neoplasm Recurrence, Local":ti, ab, kw OR "Neoplasm Metastasis":ti, ab, kw OR "Neoplasm, Residual":ti, ab, kw OR unresectable:ti, ab, kw OR advanced:ti, ab, kw OR recurrent:ti, ab, kw OR metastatic:ti, ab, kw OR "locally advanced":ti, ab, kw |
| #3 | #1 AND #2 |
| #4 | chemotherapy:ti, ab, kw OR "systemic therapy":ti, ab, kw OR "second-line":ti, ab, kw OR "salvage therapy":ti, ab, kw OR "targeted therapy":ti, ab, kw OR pharmacotherapy:ti, ab, kw OR ("immune checkpoint" NEAR/1 inhibitor*):ti, ab, kw OR ("PD-1" NEAR/1 inhibitor*):ti, ab, kw OR ("PD-L1" NEAR/1 inhibitor*):ti, ab, kw OR ("CTLA-4" NEAR/1 inhibitor*):ti, ab, kw |
| #5 | pazopanib:ti, ab OR trabectedin:ti, ab OR eribulin:ti, ab OR gemcitabine:ti, ab OR docetaxel:ti, ab OR doxorubicin:ti, ab OR ifosfamide:ti, ab OR nivolumab:ti, ab OR pembrolizumab:ti, ab OR atezolizumab:ti, ab OR avelumab:ti, ab OR durvalumab:ti, ab OR ipilimumab:ti, ab OR olaratumab:ti, ab OR brigimadlin:ti, ab |
| #6 | #4 AND #5 |
| #7 | #3 AND #6 |

### 5.3. Search strategy for Embase via ProQuest

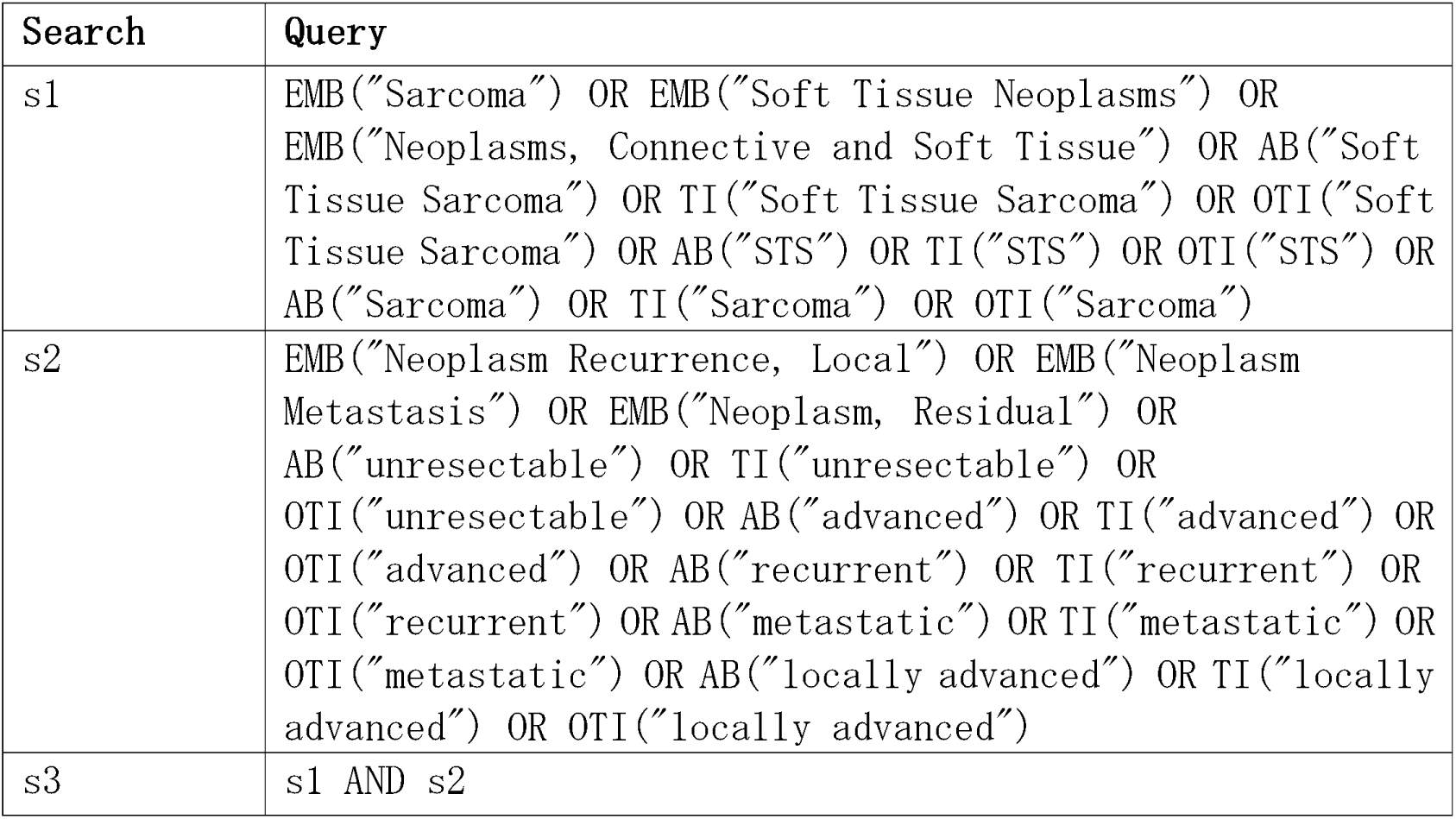
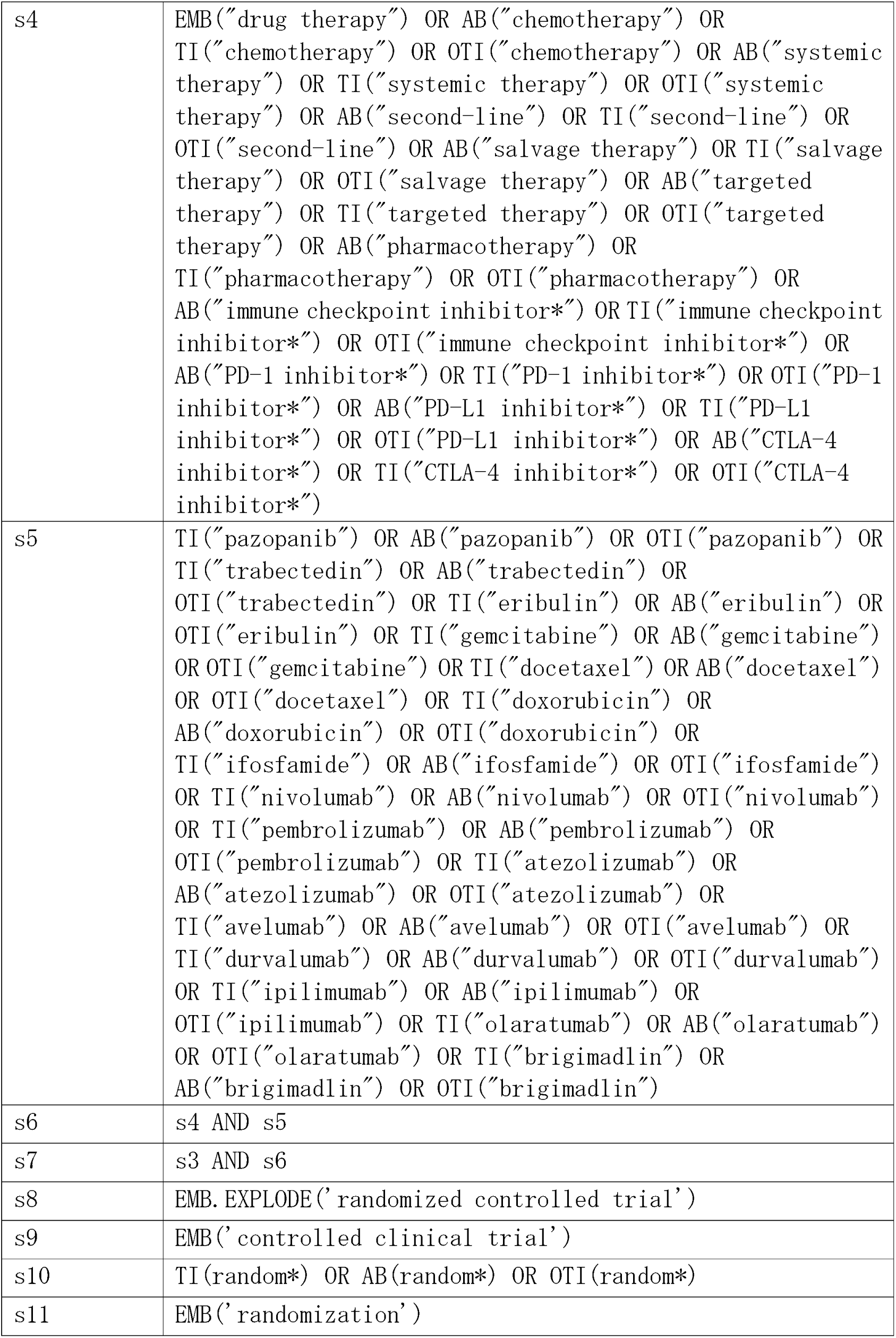
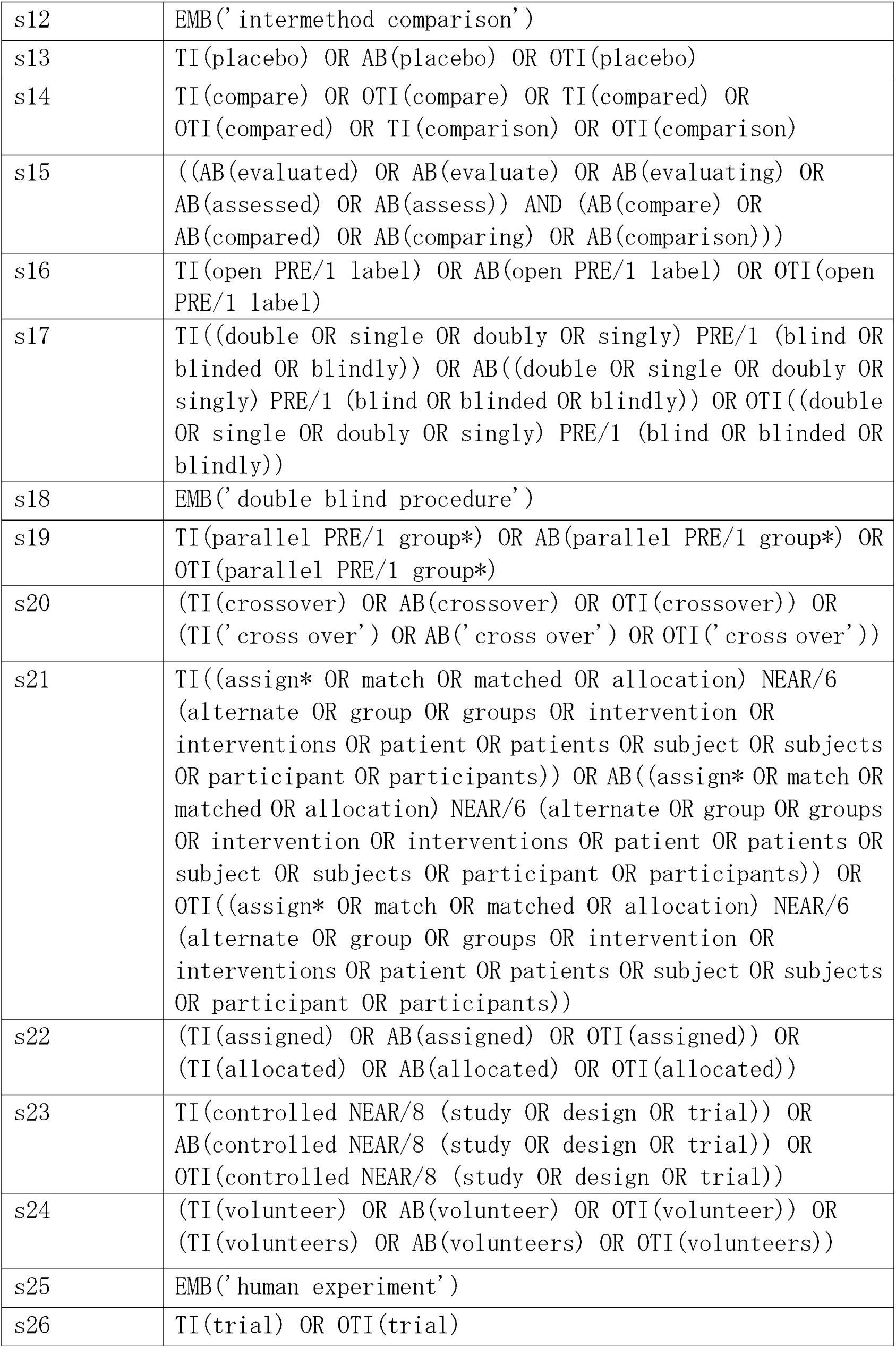
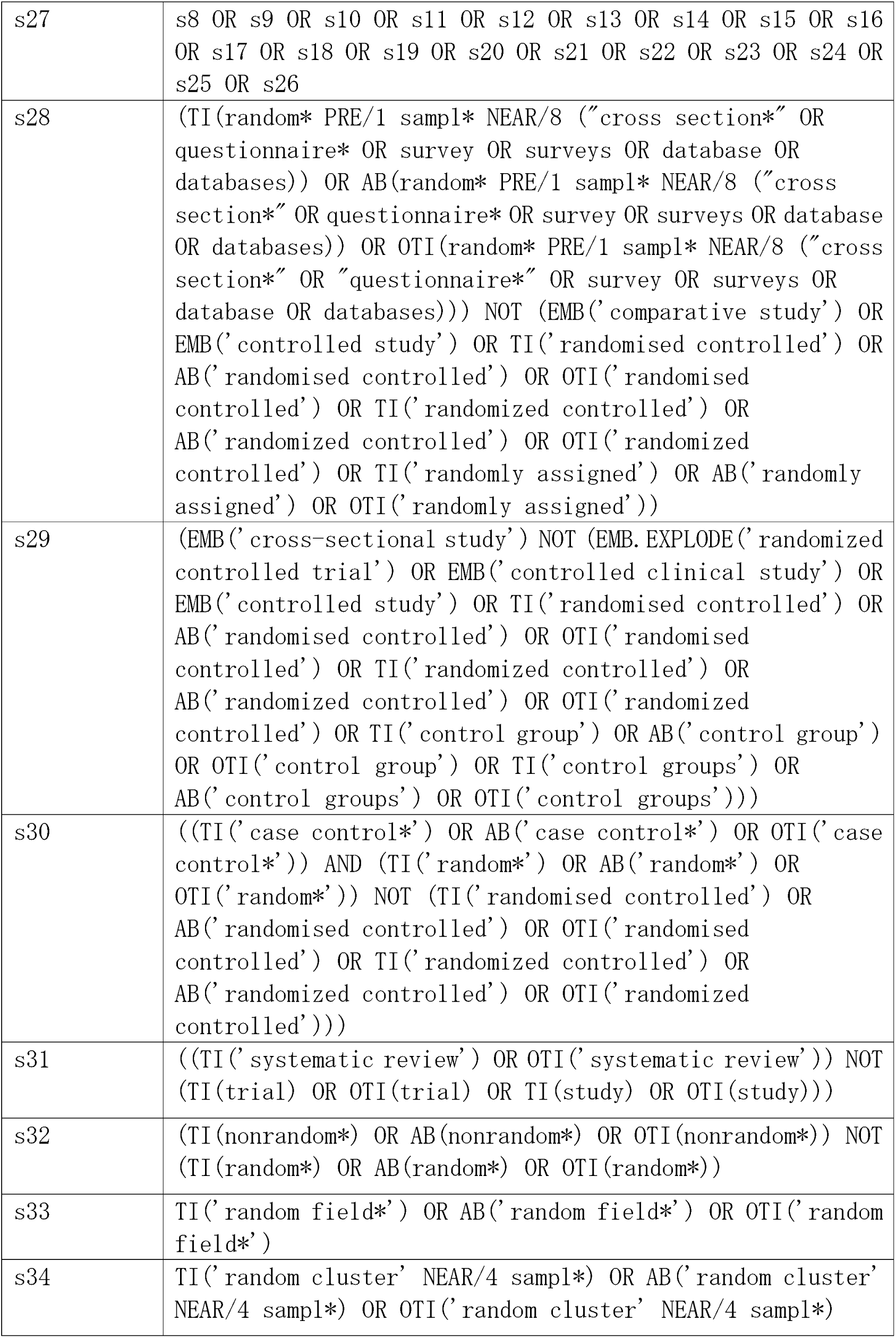
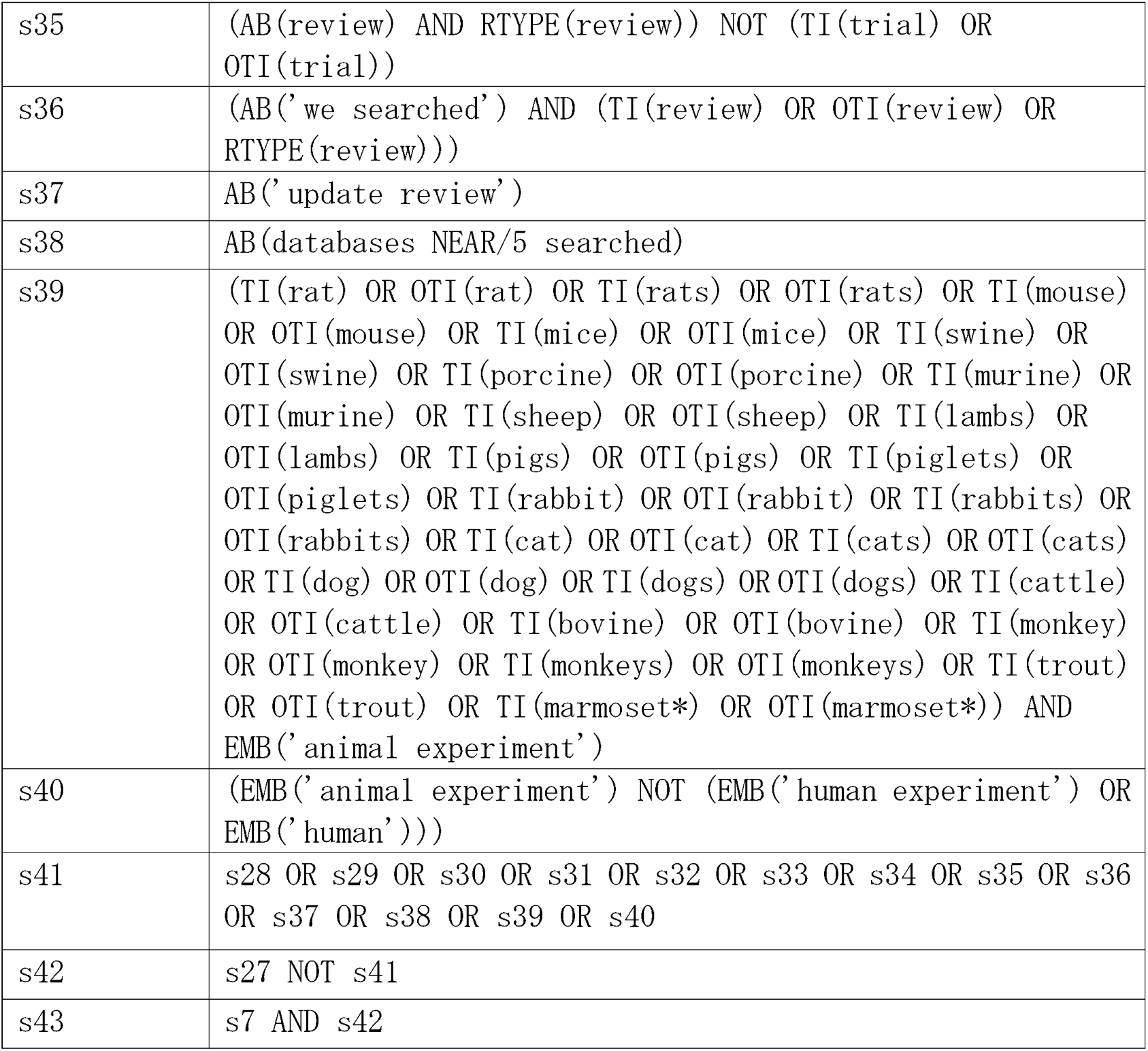

### 5.4. Search Strategy for ClinicalTrials.gov

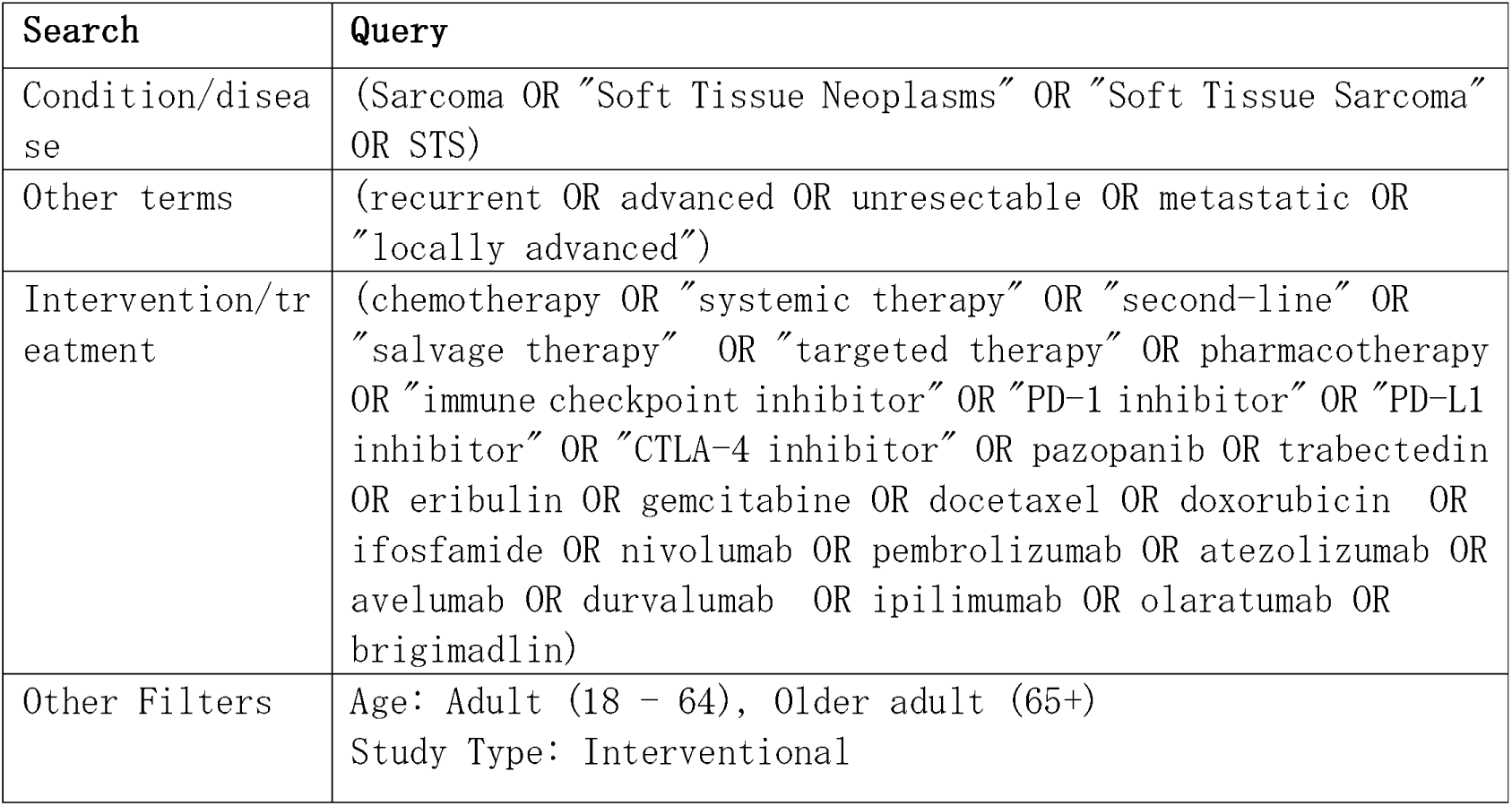

### 5.5. Search Strategy for WHO/ICTRP

| Search | Query |
| --- | --- |
| Title | NA |
| Condition | (Sarcoma OR "Soft Tissue Neoplasms" OR "Soft Tissue Sarcoma" OR STS) |
| Intervention | (chemotherapy OR "systemic therapy" OR "targeted therapy" OR "immune checkpoint inhibitor" OR pazopanib OR trabectedin OR eribulin<br>OR gemcitabine OR docetaxel OR doxorubicin OR ifosfamide<br>OR nivolumab OR pembrolizumab OR atezolizumab OR avelumab<br>OR durvalumab OR ipilimumab OR olaratumab OR brigimadlin) |
| Recruitment status | ALL |
| Phases | Phase2, Phase3 |

### 5.6. List of Abbreviations

**5.6.List of Abbreviations**
| Abbreviations | Full spell |
| --- | --- |
| CENTRAL | Cochrane Central Register of Controlled Trials |
| GIST | Gastrointestinal Stromal Tumor |
| GRADE | Grading of Recommendations Assessment, Development, and Evaluation |
| ICTRP | WHO International Clinical Trials Registry Platform |
| NBIB | National Library of Medicine Bibliographic format |
| NMA | Network Meta-Analysis |
| NCCN | National Comprehensive Cancer Network |
| ORR | Overall Response Rate |
| OS | Overall Survival |
| PFS | Progression Free Survival |
| PRISMA | Preferred Reporting Items for Systematic reviews and Meta-Analyses |
| PRISMA-P | PRISMA-Protocols |
| PRISMA-NMA | The PRISMA for network meta-analyses (NMA) extension |
| REML | Restricted Maximum Likelihood |
| RIS | Research Information Systems format |
| RCT | Randomized Controlled Trial |
| SR | Systematic Review |
| STS | Soft Tissue Sarcoma |

