## Supplementary material for "Effectiveness of Systemic Treatments in Patients with Unresectable, Advanced, or Recurrent Soft Tissue Sarcomas Previously Treated with Anthracycline-Based Therapy: A Systematic Review and Network Meta-Analysis": Version history_20260719

Date created: 19 July 2026

| Study title | Effectiveness of Systemic Treatments in Patients with Unresectable, Advanced, or Recurrent Soft Tissue Sarcomas Previously Treated with Anthracycline-Based Therapy: A Systematic Review and Network Meta-Analysis |
| --- | --- |

**Protocol (v1.3 → v1.4)**

| Subject | Before revision  (Version 1.3, May 13, 2026) | After revision  (Version 1.4, July 19, 2026) | Reason for change |
| --- | --- | --- | --- |
| 2. Objectives  2.2.3 Histology-Specific OS | Subtypes will be classified into six groups: the five most common histologies and all others combined. | Subtypes will be classified into up to five groups, consisting of the most common histologies independently reported across included trials. | Data extraction indicated that a fixed six-group classification, including a combined "others" category, does not reflect the actual distribution of reported histological subtypes across trials. The "others" category represents a clinically heterogeneous residual group that limits meaningful interpretation, and was therefore removed. |
| 4. METHODS  4.8. Subgroups and sensitivity analysis | Not specified | As a sensitivity analysis, treatment nodes may be redefined as follows: agents specifically approved for STS (e.g., pazopanib, trabectedin, eribulin) will be retained as independent nodes; combination regimens (e.g., gemcitabine plus docetaxel) will also be retained as independent nodes; and remaining monotherapy agents without STS-specific approval (e.g., dacarbazine, ifosfamide) will be consolidated into a single "traditional chemotherapy" node. | Treatment nodes were reclassified using three criteria: STS-approved agents (e.g., pazopanib) and other combination regimens (e.g., gemcitabine plus docetaxel) were retained as independent nodes, given their distinct regulatory and therapeutic identity; remaining monotherapy agents without STS-specific approval were consolidated into a "traditional chemotherapy" node as mutually substitutable options. This sensitivity analysis evaluates robustness under a more interpretable network structure, without altering the primary analysis. |
| 4. METHODS  4.9. Assessment of transitivity and inconsistency | Not specified | Trials enrolling patients with a single histological subtype will be included in histology-specific analyses only and excluded from the primary analyses of the overall population. | During data extraction, several eligible trials were found to enroll only a single histological subtype. Including these trials in the primary analysis of the overall population would violate the transitivity assumption due to systematic differences in patient population composition across comparisons. |

**Protocol (v1.2 → v1.3)**

| Subject | Before revision (Version 1. 2, February 14, 2026) | After revision (Version 1.3, May 13, 2026) | Reason for change |
| --- | --- | --- | --- |
| 2. Objectives/  3. PICO 3.4. Outcomes | LRFI and DRFI included as secondary outcomes | **Removed** (See Reason for change) | No eligible trials reported LRFI or DRFI following data extraction. These endpoints were originally derived from perioperative studies and are not applicable to the present network. |
| 4. METHODS  4.8. Subgroups and sensitivity analysis | Not specified | Sensitivity analysis restricting treatment discontinuation analysis to discontinuation due to toxicity, where data are available. | Heterogeneity in reporting treatment discontinuation (e.g., PD-related or AE-related only) was identified during data extraction. A sensitivity analysis was added accordingly. |

**Protocol (v1.1 → v1.2)**

| Subject | Before revision (Version 1.1, November 3, 2025) | After revision (Version 1.2, February 14, 2026) | Reason for change |
| --- | --- | --- | --- |
| Study Title | Effectiveness of Second-Line Treatments for Unresectable Advanced or Recurrent Soft Tissue Sarcomas: A Systematic Review and Network Meta-Analysis | Effectiveness of Systemic Treatments in Patients with Unresectable, Advanced, or Recurrent Soft Tissue Sarcomas Previously Treated with Anthracycline-Based Therapy: A Systematic Review and Network Meta-Analysis | Alignment with eligibility criteria |
| 2. Objectives | To identify effective second-line pharmacological treatment regimens from the perspective of disease control in patients with unresectable, advanced, or recurrent soft tissue sarcoma previously treated with anthracycline-based chemotherapy. | To identify effective second-line or later pharmacological treatment regimens from the perspective of disease control in patients with unresectable, advanced, or recurrent soft tissue sarcoma previously treated with anthracycline-based therapy. | Alignment with PICO |
| 3. PICO  3.1. Participants | - Aged 18 years or older | - Aged 15 years or older | Expanded age eligibility to include adolescents (≥15 years) to reflect the age ranges used in relevant clinical trials. |
| 3. PICO  3.5. Exclusion Criteria | - Trials including patients with osteosarcoma*, chondrosarcoma*, plasmacytoma, chordoma, Ewing sarcoma, rhabdomyosarcoma, Kaposi’s sarcoma, or gastrointestinal stromal tumor (GIST), carcinosarcoma  (*except for extraskeletal osteosarcoma and extraskeletal myxoid chondrosarcoma, which will be included) | - Trials including patients with osteosarcoma*, chondrosarcoma*, plasmacytoma, chordoma, Ewing sarcoma, rhabdomyosarcoma, Kaposi’s sarcoma, gastrointestinal stromal tumor (GIST), or carcinosarcoma  (*except for extraskeletal osteosarcoma and extraskeletal myxoid chondrosarcoma, which will be included) | Editorial revision to ensure correct parallel structure and avoid ambiguity in the exclusion criteria list; no substantive change to eligibility criteria. |

**Protocol (v1.0 → v1.1)**

| Subject | Before revision (Version 1.0, September 28, 2025) | After revision (Version 1.1, November 3, 2025) | Reason for change |
| --- | --- | --- | --- |
| 3. PICO  3.1. Participants | -　Patients with unresectable,　advanced, or recurrent malignant soft tissue sarcoma | -　Patients with unresectable,　advanced, or recurrent soft tissue sarcoma | Terminology clarified to ensure consistency between inclusion and exclusion criteria. |
| 3. PICO  3.5. Exclusion Criteria | - Trials including patients with osteosarcoma*, chondrosarcoma*, plasmacytoma, chordoma, Ewing sarcoma, rhabdomyosarcoma, Kaposi’s sarcoma, or gastrointestinal stromal tumor (GIST) | - Trials including patients with osteosarcoma*, chondrosarcoma*, plasmacytoma, chordoma, Ewing sarcoma, rhabdomyosarcoma, Kaposi’s sarcoma, or gastrointestinal stromal tumor (GIST), carcinosarcoma | Added carcinosarcoma to the exclusion list for clearer specification of target patient populations. |
| 3. PICO  3.5. Exclusion Criteria | Not specified in Version 1.0 | - Trials in which different routes, schedules, or doses are compared within the same drug regimen | Added to clarify exclusion of studies comparing administration methods within the same drug regimen. |
| 4. METHODS  4.2. Search Methods for identification of studies | - Embase (via Elsevir) | - Embase (via ProQuest) | Revised to accurately reflect the actual database access platform used (ProQuest instead of Elsevier). |
| 4. METHODS  4.4. Data extraction and management | Not specified in Version 1.0 | In cases where multiple reports originate from the same study, data will be handled as follows:  For studies with multiple publications (e.g., interim and final reports), time-to-event outcomes will be extracted from the report with the longest follow-up, while additional outcomes not included in the final report may be taken from other publications. Outcome-specific source references will be recorded. | Added a predefined rule for handling multiple publications of the same study to ensure consistency and transparency in data extraction. |
| 5. Appendix | 5.3. Search strategy for EMBASE via Ovid | 5.3. Search strategy for Embase via ProQuest | Revised to accurately reflect the actual database access platform used (ProQuest instead of Ovid). |
